# A small-scale Development Impact Bond for hepatitis C diagnosis and treatment in Cameroon: the way to elimination?

**DOI:** 10.1101/2023.03.24.23287688

**Authors:** C.M. Dieteren, A.C. Boers, W. Thomas, O. Njoya, R.A. Coutinho, the HEP C-IMPACT team:, T Mossus, F Essomb, G Wafeu, B Agnouanang

## Abstract

In the absence of sufficient (tax-based) healthcare financing in low-and middle-income countries, innovative financing models are needed. This study assessed quantitatively and qualitative the feasibility of a Development Impact Bond (DIB) for hepatitis C Virus (HCV) diagnosis and treatment in Cameroon. A revolving fund of up to €230,000 was made available by the investor. The outcome payor repaid the investor only in case of good performance, defined as cured patients (HCV-RNA negative). Identified HCV carriers were referred for treatment and tested for cure 12 weeks after completion of treatment, the outcome being validated by an independent party. The evaluation was guided by a recognized framework, involving interviews with relevant stakeholders (N= 22). In total, 253 (98%) patients completed treatment of which 244 (96%) are cured at week 24. We estimated that the average per patient *outcome payment* for HCV diagnosis and treatment is €1,542 and the *average costs per treated patient* is €1,858. The investor was fully repaid including the agreed interest and bonus rate. The interviews confirmed the feasibility of the DIB in a low-resource setting. This study demonstrates that a DIB can be a suitable financing mechanism for HCV services, supporting the path towards elimination. When governments do not have sufficient resources to fund such elimination programs upfront, such public-private partnerships can offer a solution.

**Highlights:**

- The availability of a short-course highly effective hepatitis C treatment paves the way towards its elimination.
- We demonstrate feasibility of the first Development Impact Bond (DIB) as a financing mechanism for hepatitis C diagnosis and treatment.
- This study shows that with an average per patient outcome payment of €1,542, one HCV patient can be diagnosed and treated in a lower-middle income country.

## Introduction

Worldwide about 58 million people carry chronic hepatitis C Virus (HCV). An estimated 290,000 of these carriers die each year from liver cirrhosis and primary liver cancer, mostly because they are unaware of their infection (World Health Organization, 2022). Therefore, the WHO goal of eliminating HCV by 2030 poses a considerable challenge. The availability and success of short-course, highly effective Direct Acting Antivirals (DAAs) with only few side effects has resulted in the recommendation for a treat-all approach (World Health Organization, 2018a). Although such a strategy for HCV elimination is cost-effective and cost-saving, most low- and middle-income countries (LMICs) with a considerable HCV burden, like Cameroon, are currently not investing in elimination (Pedrana et al., 2021).

Recently, sharp drops in DAA prices have been reached through tiered pricing, which uses a country’s national income and voluntary license agreements to determine prices (World Health Organization, 2018b). The government of Cameroon – a Central African country with about 28 million inhabitants and a high HCV prevalence especially among those aged 45 and above – also negotiated considerably lower prices for both branded and generic WHO prequalified DAA. However, because of a lack of healthcare insurance, patients are forced to pay out-of-pocket for the DAA medication as well as for diagnostics and specialist consultations. These combined costs are unaffordable for most chronic HCV carriers in Cameroon. Another barrier for early treatment is the lack of screening programs to detect asymptomatic HCV patients. Having showed high cure rates for HCV carriers with DAAs in clinical settings in Cameroon (Coyer et al., 2020), accessibility and provision of the HCV care continuum should subsequently be addressed to ensure effective large-scale implementation and potential elimination.

The unaffordability of DAA treatment for most chronic HCV carriers in Sub Saharan Africa stimulates policy makers and scientists to come up with innovative financing mechanisms. Many African public health systems fail in the delivery of efficient healthcare, partly due to a lack of (tax-based) resources, forcing many patients to pay out of pocket for the health services they use. While investing in the healthcare supply chain is often considered too risky from a private investment point of view, there are other areas of collaboration between the government and the private sector – so called public private partnerships (PPP) - increasingly happen in the healthcare sector. In many LMICs, the gap between healthcare needs and healthcare provision is the result of poor drugs supply, scarce resources, and weak healthcare infrastructures. Hence, PPP can enable vulnerable populations to access health care (Montagu et al., 2011) because it increases the budget that is available within the health sector.

A Development Impact Bond (DIB) is such an innovative financing mechanism in which pre-payment of development program expenses is provided by private investors while public agencies or donors repay the investor’s investment plus a reasonable interest rate once the program succeeds in delivering independently measurable results that are contractually agreed upon. A DIB can especially be used in healthcare when the results of a given intervention are unequivocally measurable through a qualified third party, and where there are potential governance issues. DIBs have attracted considerable interest but are relatively new and the evidence base is limited (Fraser et al., 2018).

DIBs generally involve six agents as displayed in Figure 1 (Drew & Clist, 2015). The investor provides the initial funding for the project which is directed to the intermediary. The intermediary uses this money to pay the service providers, who interact with the target population. Once the target population is reached and treated, the independent evaluator will validate the reported outcomes. The outcome payer only disburses funds to the investor based on achieved successes as reported by the evaluator.

**Figure 1.**
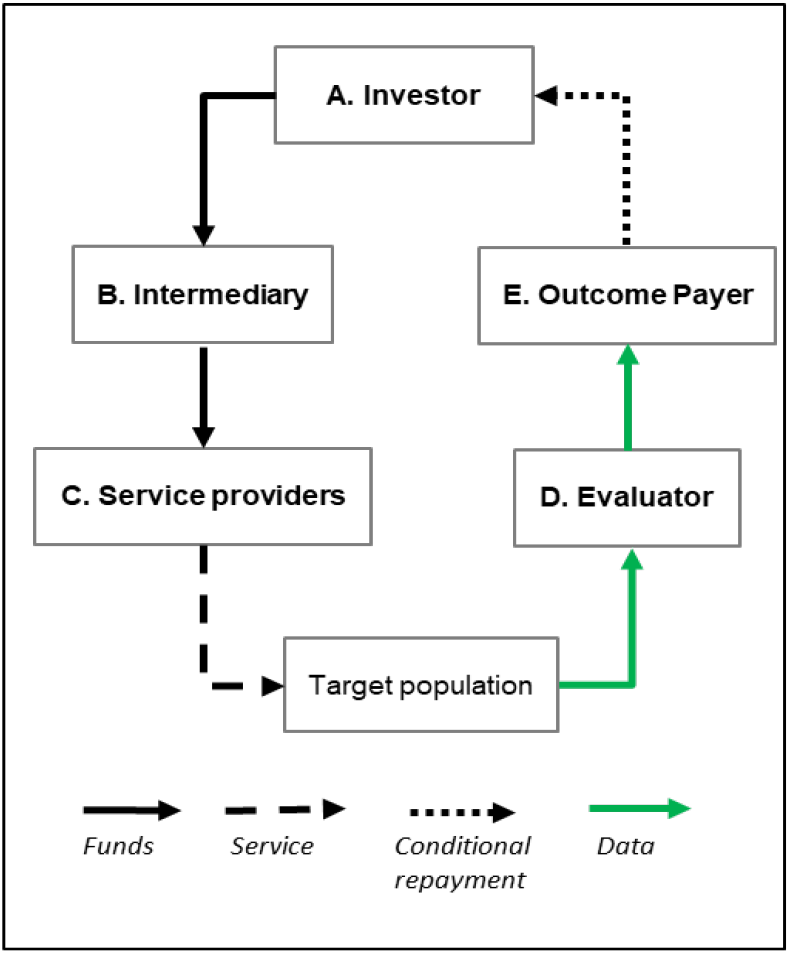
The six-agents model for DIBs.

DIBs are relatively new. Currently there are 16 of them active worldwide, of which most are implemented in Africa in the field of social welfare, employment and health (Gustafsson-Wright et al., 2022). The outcome-payer can either be the government or a donor. A DIB is generally characterized by four preconditions: i) there is a measurable outcome that can be validated by an independent verifier, ii) there is a reasonable time horizon to achieve the desired outcomes, iii) there is evidence that the outcomes can be achieved successfully and iv) appropriate legal and political conditions are in place to support the bond (Belt et al., 2017). The fourth precondition is most important when the government is the outcome payer while the third one is crucial to motivate investors to provide the pre-investment.

The reported high DAA cure rate of more than 95% over a short treatment period of 12 weeks for chronic HCV carriers suggests that a DIB might be a fitting route to generate investments for treatment programs. Hence, investing in a DIB for HCV diagnosis and treatment is likely to be an attractive way of investing for private (social) investors. Cameroon is one of the first sub-Saharan countries with a reported high cure rate (>96%) with DAA, meanwhile, the national HCV strategy to determine HCV prevalence in Cameroon is not sufficiently advanced to permit an immediate implementation of a nationwide diagnosis and treatment program. Therefore, we aim to assess the feasibility of a DIB for HCV diagnosis and treatment in the Cameroonian setting, by evaluating the execution and the financial and medical results at a small-scale. This will help identifying barriers and facilitators for future DIBs and brings in concrete information with respect to future scaling.

## Materials and methods

### Study design

We designed a prospective longitudinal study. The patient enrollment period was six months and data (on treatment status and background characteristics of each patient) was collected at the following time points: i) the start (t = 0), ii) after the treatment period (t = 12 weeks) and iii) after the 12 weeks waiting period before final testing for cure (t = 24 weeks). For HCV positives who fail on standard treatment and receive second line treatment, data was collected at t = 36 weeks and additional testing for cure took place at t = 48 weeks. The standard treatment for HCV positive patients is a fixed dose of generic pangenotypic DAA (Sofosbuvir and Velpatasvir), second line treatment is carried out with branded Sofosbuvir/Velpatasvir/Voxilaprevir. Both treatments are funded through the DIB.

### Study setting & study population

The study was carried out in Yaoundé, the capital of Cameroon in Central Africa. Participants for this study are identified through blood banks because here they do standard testing of potential blood donors for HCV antibodies to avoid transmission to patients. In the Cameroonian system, those who test positive are refused as a blood donor and are informed about being antibody HCV positive and advised to visit a medical doctor. For this study, those antibody HCV positives are referred to program clinics and assessed for eligibility for HCV treatment. As most antibody HCV positive potential blood donors did not show up at the assigned clinics, we decided to also include HCV infected persons identified by private or public Health Centers in Yaoundé. Identified eligible HCV infected persons willing to participate are treated in five clinics (University Teaching Hospital of Yaoundé, Essos Medical Center, Centre Medical la Cathedral, Central Hospital of Yaoundé and General Hospital of Yaoundé).

### Sample size justification

Based on the financial investment in the DIB, this study intended to treat 300 patients. Based on the rate of HCV positive persons identified per year in the blood banks, the estimation of the recruitment would be completed within six months. The target was to identify around 500 antibody-HCV positive donors to enroll the planned 300 HCV infected persons. Of the 500 antibody-HCV positive donors we assumed that 25% would have cleared the virus by themselves (i.e. antibody-HCV positive and HCV-RNA negative) (Aisyah et al., 2018), 10% would be false positive (first anti-HCV testing in blood bank positive but not confirmed in second test) and about 10% lost to follow-up. The final two assumptions are based on experts’ consultations.

### DIB Design

Table 1 contains characteristics of the financing mechanism. The DIB is structured as a revolving credit facility, funded by the investor (Joep Lange Institute – a Dutch NGO, www.joeplangeinstitute.org). The investor bore the social and operational risk of project success, in exchange for a basic interest rate of 5% per annum and a 1% facility fee. Success is defined as the number of chronic HCV carriers who are HCV-RNA negative at t = 24 weeks (12 weeks after completion of DAA treatment). The Agence Nationale de Recherche sur le Sida (ANRS) in Cameroon, an internationally recognized medical research and monitoring organization acts as validating agency and verifies the cured patient data. The outcome is dichotomous: cured (HCV-RNA negative) or not cured (HCV-RNA positive). Once validated, fixed outcome payments per cured patients are made by the outcome payer (Achmea Foundation: www.achmea.nl/foundation) to the investor. The investor earns bonus interest from the outcome payor of 3% per annum if the cure rate is 90% or more. The staff involved in providing HCV treatment to the target population, including doctors, nurses, pharmacists, and the coordination team, are eligible to receive a bonus of up to 10% of their annual fee. The amount actually paid is determined by the team lead based on their performance, identified as patient satisfaction and quality of data administration. The staff was aware that they could receive a bonus, but the exact amount was not communicated in advance. The target population, HCV chronic carriers, were aware that they are treated as part of the study and provided written informed consent.

**Table 1.**
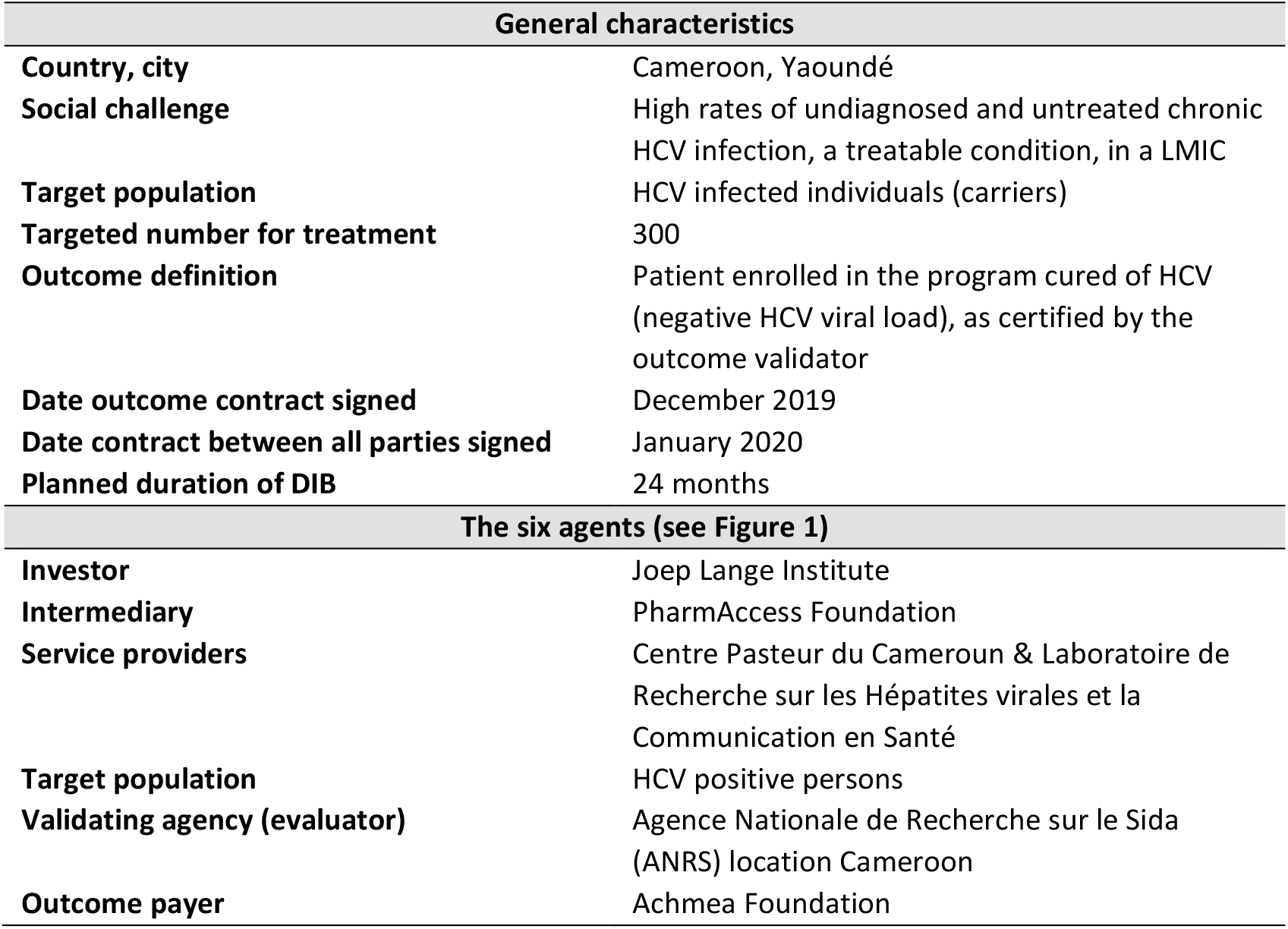
Description of the HCV Development Impact Bond.

| General characteristics |  |
| --- | --- |
| **Country, city** | Cameroon, Yaoundé |
| **Social challenge** | High rates of undiagnosed and untreated chronic HCV infection, a treatable condition, in a LMIC |
| **Target population** | HCV infected individuals (carriers) |
| **Targeted number for treatment** | 300 |
| **Outcome definition** | Patient enrolled in the program cured of HCV (negative HCV viral load), as certified by the outcome validator |
| **Date outcome contract signed** | December 2019 |
| **Date contract between all parties signed** | January 2020 |
| **Planned duration of DIB** | 24 months |
| The six agents (see Figure 1) |  |
| **Investor** | Joep Lange Institute |
| **Intermediary** | PharmAccess Foundation |
| **Service providers** | Centre Pasteur du Cameroun & Laboratoire de Recherche sur les Hépatites virales et la Communication en Santé |
| **Target population** | HCV positive persons |
| **Validating agency (evaluator)** | Agence Nationale de Recherche sur le Sida (ANRS) location Cameroon |
| **Outcome payer** | Achmea Foundation |

### Cost estimations

The treatment and diagnostic costs per patient were estimated through a financial model. In this model we considered the expected enrollment rate, adherence rate and cure rate. The costs were projected based on experience from the previous phase of this study, and on negotiated quotations for service and commodity providers. Cameroonian institutions that participated in the project (e.g. blood banks and treatment centers) were paid a fixed monthly amount for periods during which they were active. Fixed unit costs were agreed with the Centre Pasteur du Cameroon for diagnostic and analytical services, although principal reagents for viral load testing were sourced directly from the supplier. Prices largely remained fixed for the duration of the project, with only minor changes that reflected changing market conditions. To ensure adequate funding, a 5% cost contingency allowance was included in the DIB costing. A summary of the estimated cost breakdown based on the financial model – a scenario prediction and the actual costs - used can be found in Appendix 1.

We requested a patient contribution (CFA 50.000 / €76), less than one fourth of the negotiated price of the treatment medication, to ensure buy-in and to further motivate the patients to adhere to treatment. If patients were unable to pay the own contribution, they were not excluded from the study. Based on the patient contribution and overall forecast costs, we defined the funds necessary from the investor to fully finance the project. Transparency in costs was needed to determine the size of the cash-flows (the arrows in Figure 1). The investor (Joep Lange Institute) committed a short-term, revolving investment of up to a maximum outstanding of €230,000. This commitment was based on the estimated maximum outstanding investment per the financial model underlying the DIB. The maximum potential outcome payment for the outcome payer (Achmea Foundation) was planned at €397,800 based on outcome payments of €1,326 per cured patients, to a maximum of 300 cured patients. The outcome payments were expected to fully reimburse the investor as well as to cover the basic and bonus interest on the investment and fee to use the facility.

### Evaluation framework

This evaluation study is guided by the six agents model as depicted in Figure 1. In our setting, an experimental design with an intervention and comparable control group is not possible and thus we cannot claim to estimate effects. However, we evaluated the DIB in a systemic manner by means of the proposed framework (Appendix 2). This framework is organized around a theory of change showing how inputs into DIBs lead to processes which are expected to produce impacts. The evaluation consists of qualitative and quantitative components.

#### Qualitative component

We identified relevant actors for the interviews based on the six-agents model. The interviews were guided by the framework presented in Appendix 2. Additionally, we conducted interviews with relevant contextual actors (the Cameroonian government) and experts in the field of health financing. Interviews were conducted both in-person and online. The interviews took place in a neutral environment where the respondents felt safe and comfortable. Before the start of the interviews, respondents were asked to read and sign the informed consent form. All interviews were recorded and transcribed. The transcripts were analyzed in the qualitative analysis software NVivo. The six assumptions and the four processes set out in the framework (Appendix 2) were applied in the thematic coding analyses of the transcripts.

#### Quantitative component

We visualized the entire enrollment and treatment procedure to generate insights in the lost to follow-up, drop-out and cure rates. Data collection on background characteristics of those that start treatment included: gender, age, marital status, employment status, poverty probability index (PPI) score (Schreiner, 2016) and HIV-status. The Poverty Probability Index (PPI) tool consists of ten low-cost questions that were developed as part of Cameroon’s 2014 Household Survey to estimate the probability that a household has a consumption below a given poverty line. The PPI score can theoretically range between 0 and 100, with 0 indicating the highest poverty likelihood and 100 the lowest. To quantify the insights of the financial results of the DIB, we assessed the financial cash flows between the participating agents and estimated the per patients costs.

### Ethical approval

The study was approved by the Cameroon National Ethics Committee (2019/07/1178/CE/CNERSH/SP) and the MoH. All study staff signed a confidentiality agreement.

## Results

### Study population

Between January 2020 and July 2021, 52,307 potential blood donors were screened for antibodies-HCV at eight identified blood banks around Yaoundé. From these 52,307 individuals, 439 potential donors were anti-HCV ELISA positive, informed about their test result and referred to participating clinics of whom only 112 antibody-HCV ELISA (i.e. laboratory test) positives (26 %) were seen at the treatment clinics. Therefore, to complete required sample size an additional 264 HCV-antibody positive persons identified at general clinics were received at the treatment clinics. Of these 376 HCV-antibody positive individuals, a total of 258 chronic HCV carriers started treatment. In total 118 individuals were excluded because they were HCV-RNA negative (cleared HCV infection) or not eligible because of various reasons (see Figure 2). Out of the 258 individuals that started treatment, 253 individuals (98%) completed the treatment at week 12. At week 24, 244/253 (96%) are cured and 8 out of the 9 individuals – one person refused – who were not cured at week 24 received second line treatment for 12 weeks. All 8 were cured 12 weeks later.

**Figure 2.**
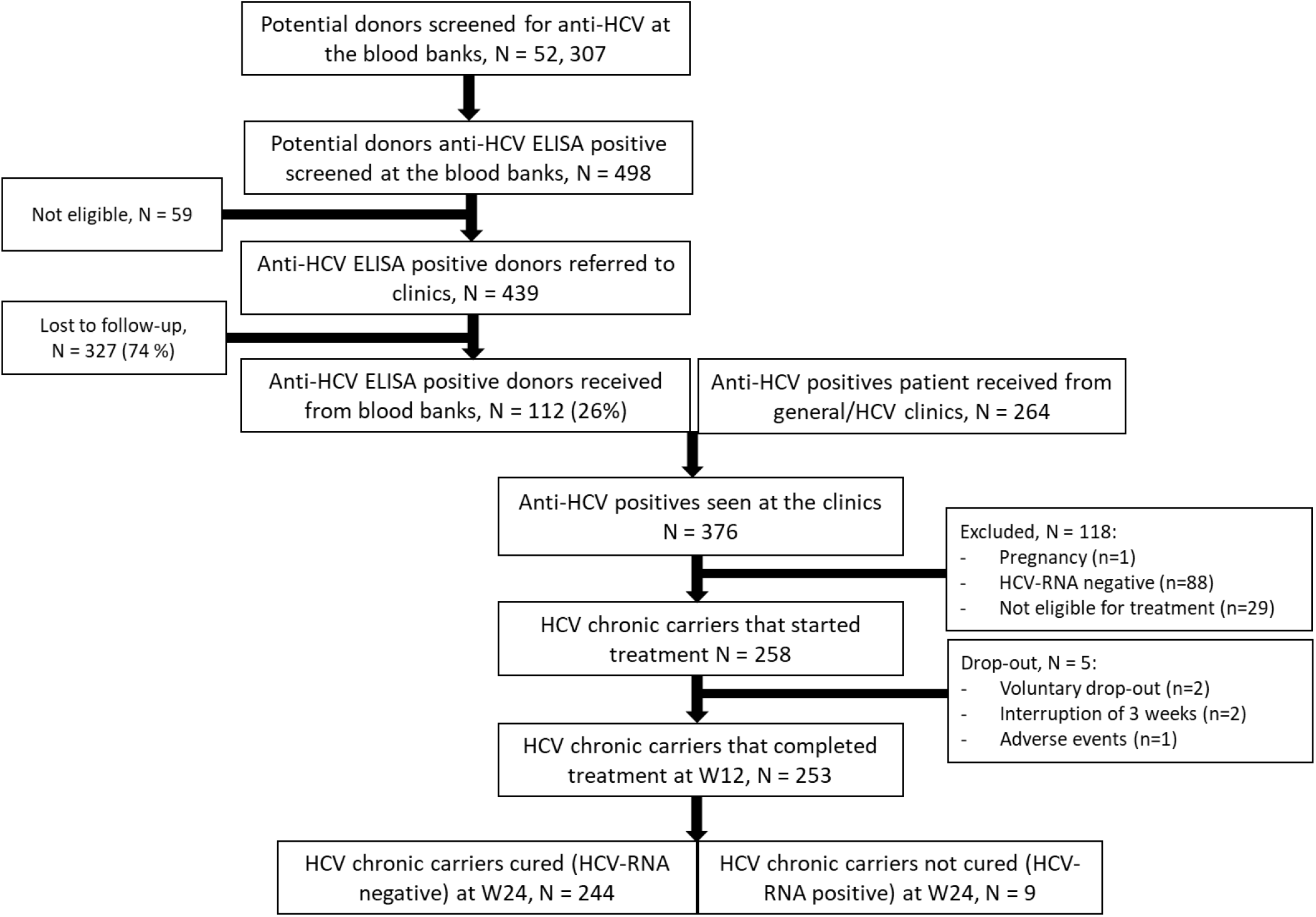
Flow chart of patient selection.

### Participants characteristics

Of the 258 participants who started treatment, 157 (61%) are female and the average age is 62 years (Table 2). The majority (53%) is married, and a quarter is widowed. One-third of the participants is unemployed, the median PPI score is 67.5 (IQR 50 – 69.2). In addition, 4% of the study population is HIV positive.

**Table 2.**
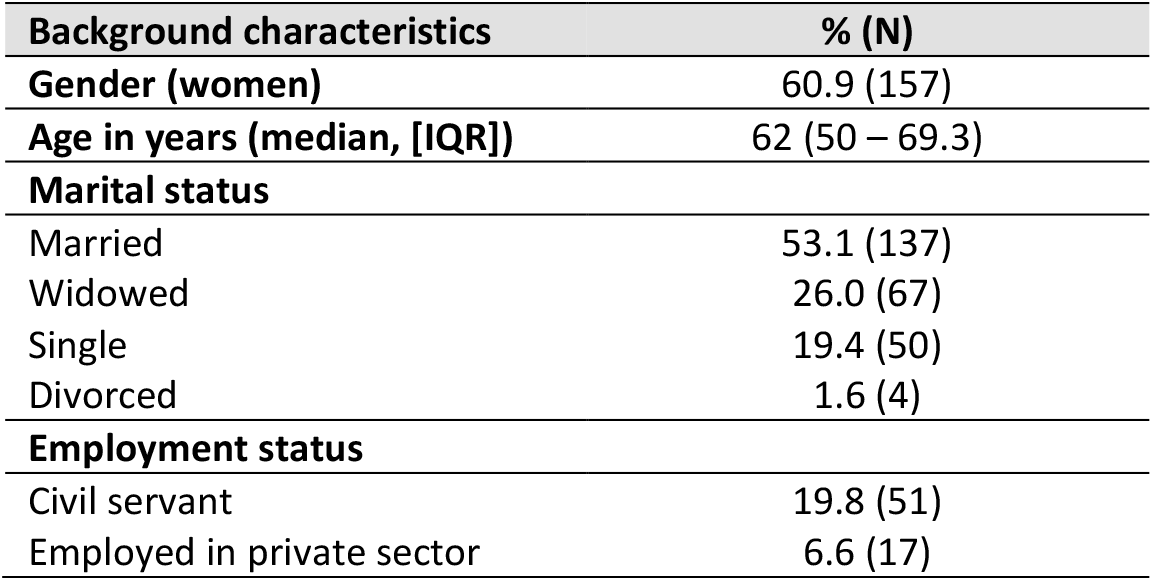
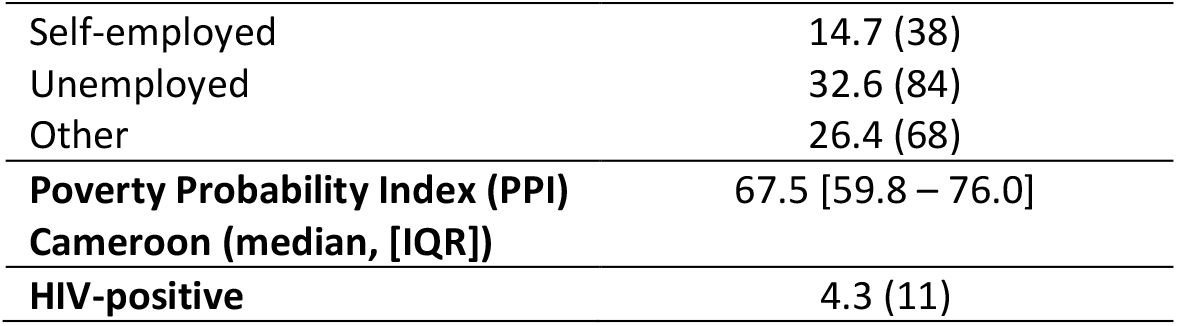
Study sample (patients that started treatment) characteristics (N=258)

### Financial outcomes DIB

The study was launched just before the start of the COVID-19 pandemic and enrollment of patients was paused subsequently for six months. To continue the project, three agents – the investor, intermediary and outcome payer – agreed to invest additional money in the DIB. The total unexpected costs due to COVID-19 were €72,500 and was paid directly into the project as a once off payment in January 2021. The unexpected costs were related to a longer recruitment period and additional staff costs. In addition, the target enrollment number was reduced from 300 to 240 because the inclusion rate was lower than anticipated.

To provide a proof of principle of the DIB mechanism, the outcome payer (Achmea Foundation) opted to redesign the financing structure whereby delays and additional costs associated with COVID-19 (e.g., extended recruitment period) were separated from the remainder of the program. This led to a split of the patient cohort and outcome payments into two separate groups. Table 3 provides an overview of the results of the financial outcomes of the DIB. Group A – the initial group – including all patients enrolled through a cut-off date set at 31 December 2021. For this group, the outcome payment per cured patient was €2,050. For the second group – Group B - the outcome payment per cured patient was reduced to €1,672. Outcome payments per cured patient were effectuated for 85 patients under Group A and 134 patients under Group B until the maximum potential outcome payment amount of €397,800 was reached. Due to this design, outcome payments were made for these 219 patients although a further 25 patients were verified as cured for which no outcome payment was released. This explains why the average outcome funding per *treated* patient - calculated as the amount of total outcome payments made (€397,800) divided by the number of patients who started treatment (n=258), equaling €1,542 per patient - is lower than either Group A or B outcome payments per cured patient. The same applies to the average outcome funding per *cured* patient (n=244), resulting in €1,630. Additionally, the outcome payments per treated and cured patient for both Group A and B are higher than the initial outcome payment per cured patient set at €1,326 because the adjusted target enrollment number was lower (n=240) than originally planned (n=300).

**Table 3.** Results of the financial commitments and average per patient costs.

| Financial commitments |  |
| --- | --- |
| Investor | Joep Lange Institute |
| Basic interest rate | 5% per annum |
| Bonus interest rate (if cure rate >90%) | 3% per annum |
| Facility fee | 1% flat |
| Total investment commitment | €230,000 |
| Actual total investment (capital recycling) | €339,574 |
| Total actual return | Total interest of €17,150<br>Fee of €2,300 |
| Outcome payer | Achmea Foundation |
| Maximum potential outcome payment | €397,800 |
| Outcome payment made | €397,800 |
| Additional funding related to COVID-19 | €72,500 |
| Total patient contribution | € 19,512 |
| Actual outcome payment per cured patient | €2,050 (first 85 patients)<br>€1,672 (thereafter) |
| Per patient costs |  |
| Average outcome funding per treated patient | €1,542 |
| Average outcome funding per cured patient | €1,630 |
| Total funding per treated patient | €1,898 |
| Total funding per cured patient | €2,007 |
| Average cost per treated patient | € 1,858 |
| Average cost per cured patient | € 1,969 |

The numbers presented in the paragraph above ignore the additional funding provided due to the delays and consequences of Covid-19 (€72,500) as well as the contributions paid by the patients. In the model it was assumed that only 90% of the patients would pay the own contribution while in reality 256 out of the 258 patients that started treatment fully paid their contribution, equaling 99% and totaling €19,512. Factoring in these two amounts, the total amount of funding provided for the treatment program equals €489,812. The funding available per patient who started treatment (n=258) thus equals €1,898, which in turn leads to €2,007 per cured patient (n=244). The average costs per patient are based on the different cost segments totaling €480,315. A stratification by all cost elements (e.g. medication costs, diagnostics costs) is presented in Appendix 1. The average costs per treated patient is € 1,858 and per cured patient is € 1,969.

### Qualitative analyses

We conducted 11 interviews, involving 22 persons (see Appendix 3). Some interviews took place in a group setting (maximum of five participants), while other interviews had one participant. Nine interviews can be categorized within the six-agent model, the other three were contextual actors. We identified common themes throughout the manuscripts and present these separately for the six agents and the contextual actors below.

#### The six-agents

##### Relationships between the agents

During the set-up of the DIB, *the investor, the intermediary* and *the outcome payor* were closely in contact with each other to develop and sign the legal contracts among those parties. The three-party contract stipulated that repayment of the facility was only to be made upon achievement of successes and provided for a revolving structure. This was a new structure for all involved parties, as most NGO’s are used to the regular way of project financing which usually involves a donation agreement in the form of a lump sum payment. Hence, all parties had to identify their role in this, and this led to an extended period of contract drafting.

The *service providers* and the *outcome evaluator* had to work closely with each other. The *outcome evaluator* visited the laboratory once every month to verify the reported results. Employers at the laboratory explained that they were used to this procedure because they also deliver results for the Demographic and Health Survey (www.dhsprogram.com). This procedure also requires a thoroughly independent evaluation and is therefore comparable in many aspects. It was noted, however, by the *outcome evaluator* that that payout for the achieved success did not happen in real time due to the multiple validation steps.

> *“We did every month the validation. So once a month I should collect all the code of people for whom I received the notification from the base that they were declared as cured. …… But once every two months I received a message from the financing agency: ‘did you really validate’, ‘can you confirm that this and this*.*’” (Respondent I)*

##### Compared to the regular way of working

The project introduced minor changes in the regular way of working of the *service providers*. To have an external data validation system, the service providers had to work with a specific data-entry system. This system was considered as trustworthy and good.

The service providers that delivered the treatment to the patients were to a certain extent incentivized to improve the quality of care, because they would receive a bonus for good performance. Service providers that arranged all data entry and checked all systems were additionally incentivized with a small bonus when the project was successfully completed, and this was motivating for them.

> “*I was already conditioned to focus on the good results. But now, it was more encouraging and stimulating because there was a bonus behind the work*.*” (Respondent II)*

##### Positive outcomes

Three main themes were extracted as positive outcomes of the DIB. Firstly, the agents agreed that the DIB has led to a fairly accurate costing estimates of HCV diagnosis and treatment. These costing insights are considered as innovative and valuable findings.

> “*We have given it a price. We can show quantitatively up to the euro what HCV diagnosis and treatment costs. [*..*] This gives transparency for a future outcome payer also if they only want to pay a certain part*.*” (Respondent III)*

Second, the DIB has led to advanced data systems. For a DIB to work, a transparent data system is required. This had to be developed and implemented. While it required training, and clinicians continued to also work with their paper files, it is also considered as a step forward into electronic patient files.

> *“I don’t know if I can state it as an effect, but the consequence of the DIB is good availability of the data*.*” (Respondent II)*
>
> *“The electronic patient file is something new. That is not common here and I think that is a good experience, although it was sometimes difficult*.*” (Respondent II)*

Third, it was mentioned that the DIB has generated additional medical insights as the same cure rates in Cameroon were achieved compared to as in high-income countries.

#### Contextual actors

##### A market for DIBs

A prerequisite for a DIB to be successful and sustainable is a flourishing DIB market. In the past decade, the DIB market did not grow as exponentially as many were hoping for. According to the interviewed experts, it is challenging to find a suitable performance indicator because there are many exogenous factors that could influence the result. As highlighted both by the six agents and the contextual actors, the performance indicator for HCV is straightforward and easily fits a DIB structure. The high cure rate, also in the Cameroonian setting, is an interesting selling point for investors as their financial risk is substantially reduced by this. In addition, it was advocated that a DIB structure would make it economically and politically easier for a government to justify their spendings.

> *“Politically it is easier for them [the government] to do because they can say to their people: we are paying for a 1,000 people this year to be cured for Hepatitis C, but we are only paying if they are cured. […] And economically, because you have confidence that you would be only paying for a most affordable price*.*” (Respondent IV)*

The above quote suggests a scenario in which the government takes over the role of outcome payor. An important note here is that because this DIB is a revolving fund and can be scaled up over time, multiple players can buy into this financing mechanism. Such a buy-in market can be attractive for the government so they can split costs among themselves and donors. More importantly, for donors to step into a larger scale DIB it was argued that the support and involvement of the government is a prerequisite since HCV is a problem in the public domain.

> *“The funding of the State is indispensable. However, these funds are always likely to be insufficient and pose a problem of mobilization*.*” (Respondent IV)*

##### Hepatitis C-virus

On the one hand, HCV as the subject of a DIB was considered as appealing due to the nature of the performance indicator (short treatment period; high cure rate), but on the other hand the number of people advocating for investments in HCV elimination is relatively small compared to other diseases (e.g., HIV/AIDS, tuberculosis and malaria).

> *“Hepatitis C is not so high on the priority agenda: middle aged people get it and it is a salient disease. So even the people that should be fighting to get the treatment, don’t know they have it. But they end up with a painful death*.*” (Respondent V)*

Thus, there are several reasons why HCV is not high on the agenda but what should be emphasized according to the interviewees is that the explicit costs of curing people from HCV are not so high and that the opportunity for this is enormous. It was advocated among many interviewees that HCV has the potential to become an attractive disease to invest in, because there are now clear cost estimates and an affordable medicine that almost guarantees cure.

## Discussion

This study describes and evaluates the successful implementation of a small-scale DIB for HCV diagnosis and treatment in Cameroon. In total, 258 chronic HCV carriers were treated with a cure rate of 96%. The DIB showed that with an average per patient outcome payment of only €1,542 it can treat one HCV patient. Worldwide, this is the first DIB devoted to HCV and the results show that the context of HCV diagnosis and treatment is suitable for this innovative financing mechanism.

The first HCV DIB is closing which marks the end of this study, while all elements are in place to benefit from the economies at scale principle. The costs of HCV service delivery used to be a black box, but this study generated valuable insights in this regard. We demonstrate that the costs to treat one chronic HCV carrier is €1,858 in a lower-middle income country. However, this cost estimate can be further optimized because it is likely to be substantially lower when assessed at scale. For instance, given the relatively small size of this study the local fixed costs were much higher. Additionally, when implemented with governmental support many existing public health human resources could be used. In combination with an accurate HCV prevalence estimate it can be estimated how much investment is required to eliminate HCV in the Cameroonian setting. Such an ambition would fit nicely in a national strategy to eliminate HCV. Such a strategy is currently missing in Cameroon. In fact, Cameroon belongs to the countries that score very low on the HCV policy score index (Palayew et al., 2020). HCV seems to lack a form of “sexiness” for donors and governments to prioritize this disease, possibly explained by the fact that the disease is mostly prevalent among middle-aged and older people (Thrift et al., 2016). Additionally, this is a silent disease: the course of the disease is asymptomatic for a long time resulting in many people being unaware of their infection and society not experiencing HCV as a danger (Thrift et al., 2016).

This study showed that a strong financial case can be made for an HCV DIB. The financing structure could be appealing for governments in LMICs since it moves the upfront costs of service delivery to investors, and it eliminates the risk of paying for services that prove to be ineffective. However, it also requires the government to create (additional) health budget in the context of ongoing dependence on donor funding. For donors this can be a great opportunity to cooperate with the government and to ensure a measurable and sustainable impact of their funding. The bundling of different stakeholders, as private investors are also included, exemplifies a typical private-public-partnership. It is recognized that to succeed in disease elimination, the public health approach should include both public and private stakeholders and facilitate cost sharing (Ward & Hinman, 2019). When upfront costs are less of an issue, it could also be suggested that the government acts as an investor, as in that role they are incented to make sure that the program is carried oud adequately which consequently leads to reduced costs and increased revenues. This, however, is likely to be more useful in high-income settings.

Parallel to the DIB central in this study, there were two other DIBs being implemented in the Cameroonian setting. The Cameroon Kangeroo Mother Care (KMC) DIB was launched late 2018 and was closed after two and a half years of service delivery. Across 10 hospitals, 1,221 babies’ received quality KMC. The other Cameroonian DIB, The Cataract Bond, is has a planned closing date in March 2023. By then, the target is to have carried out 18,000 cataract surgeries (in 5 years). While the Cameroon Ministry of Public Health is the outcome funder in the KMC DIB, the Cameroon Cataract Bond – like the HCV DIB – is fully funded by donors (Oroxom et al., 2018; Savell & Eddleston, 2021). The general acknowledgement of a DIB market by the Cameroonian government is demonstrated by the establishment of a separate unit within the Ministry of Public Health that coordinates the performance-based financing activities. Even though the KMC DIB is currently closed, the sustainability of this DIB is captured in a follow-up plan in which the collaboration with the Ministry of Public Health is key. The HCV DIB in this study was designed solely with funding from non-governmental organizations. Although there were several attempts to onboard the government in the DIB as a (partly) outcome payer, an agreement has never been made.

A large-scale HCV DIB could be implemented in a stepwise manner: the government may start with only paying a part of the outcome payment, supplemented by other payers and if feasible this amount can slowly be increased over time. An interesting by-catch of this mechanism would be that it can lead to increased trust in the government from the population. The government can justify their spending because they only pay a fixed amount for a cured patient. Conversely, this case is also interesting for private (social) investors as their risk of losing money is low and they have a high likelihood to create sustainable social impact.

A striking observation in the study is the high drop-out rate. This rate refers to the group of potential blood donors who were lost after they were identified as chronic HCV carriers and thus were eligible to receive (almost) free life-saving treatment. It could be that these individuals are not aware of the seriousness of the disease as they do not have symptoms, requiring better and more intensive counselling after diagnosis. To increase the treatment participation of chronic HCV carriers an innovative Test-and-Treat approach is recommended where carriers are treated immediately after being diagnosed with a rapid test. Such an approach has previously proven its value in HIV treatment (Gardner et al., 2011).

### Limitations

The following four limitations of the study need to be acknowledged. Firstly, this study does not allow to obtain effect estimates of this DIB intervention. That would require an RCT with three arms: i) no financing mechanism, ii) the existing financing method, and ii) the DIB financing method. We were not able to run an experimental study with a counterfactual and therefore we cannot claim to actually estimate and quantify effects. Secondly, the DIB involved some form of incentivizing the service providers by providing small bonuses based on good performance. It is considered as a missed opportunity that there were no further quality incentives incorporated in the design. Thirdly, our sample is relatively wealthy and, in that regard, not representative of the population in Cameroon. Referral from a peripheral provider to a more complex HCV treatment center automatically selects for the more affluent. However, the composition of the study sample is of less concern when assessing the feasibility of a financing mechanism. It may only explain the patient contribution rate if 99%, which was likely to be lower with a more representative sample. Finally, for continuity of the project we conclude that the government should have been involved from an earlier stage. This proved to be a challenge later in the project. The actual involvement of the government in donor funded projects is increasingly being seen as a pillar for sustainability and continuity (Theobald et al., 2018) and should be considered necessary before starting similar projects.

## Conclusions

Thanks to the availability of highly effective treatment, worldwide elimination of HCV is an achievable midterm target. When governments do not have sufficient resources to fund such elimination programs upfront, public-private partnerships can offer a solution. Our small-scale HCV DIB demonstrate that a DIB is a suitable approach for HCV diagnosis and treatment and therefore has the potential to treat chronic HCV carriers in LMICs. An important requirement for scaling and longer-term sustainability is that pertinent governments assume (partly) the role as outcome payer from the start.

## Data Availability

All data produced in the present study are available upon reasonable request to the authors.

## Acknowledgements

The authors wish to thank all HEP C-IMPACT participants and the participating clinics. In addition, they would like to thank the following persons from the study team for their support: Dr. M. Kowo (Yaoundé University Teaching Hospital), Dr. Dang Babagnack (Centre Médical la Cathédrale), Dr. P. Talla (Yaoundé General Hospital), Dr. E. (Tchoumi Yaoundé Central Hospital), Dr. D. Simo Kamto (Essos Hospital Center) and Dr. R. Njouom (Pasteur Institute). We also would like to thank Dr. Igna Bonfrer for her valuable comments to the draft manuscript.

## Classification codes

I10, I15, I18

## CRediT author statement

**C.M. Dieteren:** Investigation, Formal analysis, Writing – Original Draft, Visualization. **A.C. Boers:** Conceptualization, Methodology, Investigation, Writing – Review & Editing, Project administration. **W. Thomas:** Conceptualization, Methodology. **O. Njoya:** Conceptualization, Resources, Writing – Review & Editing. **R.A. Coutinho:** Conceptualization, Methodology, Writing – Reviewing & Editing, Supervision

