## Appendix for "A small-scale Development Impact Bond for hepatitis C diagnosis and treatment in Cameroon: the way to elimination?"

### Appendix 1. Predicted and actual costs breakdown

| | Budgeted Costs | | | Actual Costs | | |
|---|---|---|---|---|---|---|
| | At Year-end 2020 | | | Project end | | |
| | 240 Patient Scenario | | | 258 Patients Enrolled | | |
| | Per Patient Amounts in € | Total Amounts in € | Percents | Per Patient Amounts in € | Total Amounts in € | Percents |
| **Medication** | | | | | | |
| Primary treatment Medication | 345 | 82,800 | 17.9% | 357 | 92,026 | 19.2% |
| Retreatment Medication | 59 | 14,040 | 3.0% | 67 | 17,323 | 3.6% |
| **Total medication** | **€ 403.50** | **€ 96,840.00** | 21.0% | **€ 424** | **€ 109,349** | **22.8%** |
| **Diagnostic Costs** | | | | | | |
| **Viral Load Tests** | | | | | | |
| RNA Viral Load Lab Costs (S -4) | 44 | 10,448 | 2.3% | 63 | 17,015 | 3.4% |
| RNA Viral Load Lab Costs (S 24) | 16 | 3,840 | 0.8% | | | |
| RNA Viral Load GeneXpert Cartridges | 50 | 11,973 | 2.6% | 63 | 16,129 | 3.4% |
| **Total Viral Load per Enrolled Patient** | **€ 109** | **€ 26,261** | **5.7%** | **€ 125** | **€ 33,143** | **6.7%** |
| **Other Diagnostics** | | | | | | |
| ELISA | 19 | 4,473 | 1.0% | 7 | 1,745 | 0.4% |
| Basic Labs | 42 | 10,077 | 2.2% | | | |
| Tests for Complications (per enrolled) | 13 | 3,155 | 0.7% | 60 | 15,514 | 3.2% |
| Retreatment Diagnostics | 12 | 2,761 | 0.6% | | | |
| **Total Other Diagnostis** | **€ 85** | **€ 20,467** | **4.4%** | **€ 67** | **€ 17,260** | **3.6%** |
| **Total Medication & Diagnostic Costs** | **€ 598** | **€ 143,568** | **31.1%** | **€ 616** | **€ 159,752** | **33.1%** |
| **Cameroonian Medical Support Costs** | | | | | | |
| **Salaries and Site Support** | | | | | | |
| Blood Bank Teams | 163 | 39,039 | 8.4% | 78 | 20,030 | 4.2% |
| Clinical Treatment Teams | 223 | 53,543 | 11.6% | 148 | 38,268 | 8.0% |
| Project Implementation Team | 471 | 112,963 | 24.4% | 541 | 139,513 | 29.1% |
| **Total Cameroonian Salaries and Site Support** | **€ 856** | **€ 205,545** | **44.5%** | **€ 767** | **€ 197,812** | **41.3%** |
| Other Cameronian Costs (logistical & admin) | 178 | 42,661 | 9.2% | 155 | 39,889 | 8.3% |
| **Total Cameroonian Medical Support Costs** | **€ 1,034** | **€ 248,206** | **53.7%** | **€ 921** | **€ 237,700** | **49.6%** |
| **Offshore Project Support Costs** | | | | | | |
| Outcome Verification (ANRS) | 71 | 17,064 | 3.7% | 80 | 20,652 | 4.3% |
| Facility Administration (GLAS) | 104 | 25,000 | 5.4% | 116 | 30,000 | 6.3% |
| Financing Costs (Fees + Interest) | 58 | 13,838 | 3.0% | 75 | 19,450 | 4.1% |
| Electronic CRF | 60 | 14,380 | 3.1% | 49 | 12,760 | 2.7% |
| **Total Offshore Project Support Costs** | **€ 293** | **€ 70,282** | **15.2%** | **€ 321** | **€ 82,863** | **17.3%** |
| **Total Per Patient Costs** | **€ 1,925** | **€ 462,055** | **100.0%** | **€ 1,858** | **€ 480,315** | **100.0%** |
| **Funding** | | | | | | |
| Patient contribution | 69 | 16,463 | 3% | 76 | 19,512 | 4% |
| Outcome payments | 1,658 | 397,800 | 82% | 1,542 | 397,800 | 81% |
| Additional COVID-19 funding | 302 | 72,500 | 15% | 281 | 72,500 | 15% |
| **Total funding available** | **€ 2,028** | **€ 486,763** | **100%** | **€ 1,898** | **€ 489,812** | **100%** |
| **Contingency** | **€ 103** | **€ 24,709** | **5.35%** | **€ 37** | **€ 9,497** | **1.98%** |

**Notes.** The actual total per patient cost (€1,858) is lower than the modelled total per patient cost (€1,925) because of efficiencies of scale generated through more patients having been included in treatment (n=258) than was estimated in this prediction model (n=240). For this same reason, actual total program costs (€480,315) were higher than modelled total costs (€462,055). The higher costs could be accommodated due to the 5% funding contingency allowed for in the model. During the course of the program, actual costs were regularly checked against expected expenditure to ensure that the program did not exceed the budget. At the end of the program, available funding exceeded actual costs by around €9,500, demonstrating that the applied financial model functioned accurately.

#### Appendix 2. Framework for evaluating Development Impact Bonds, based on proposal DFID

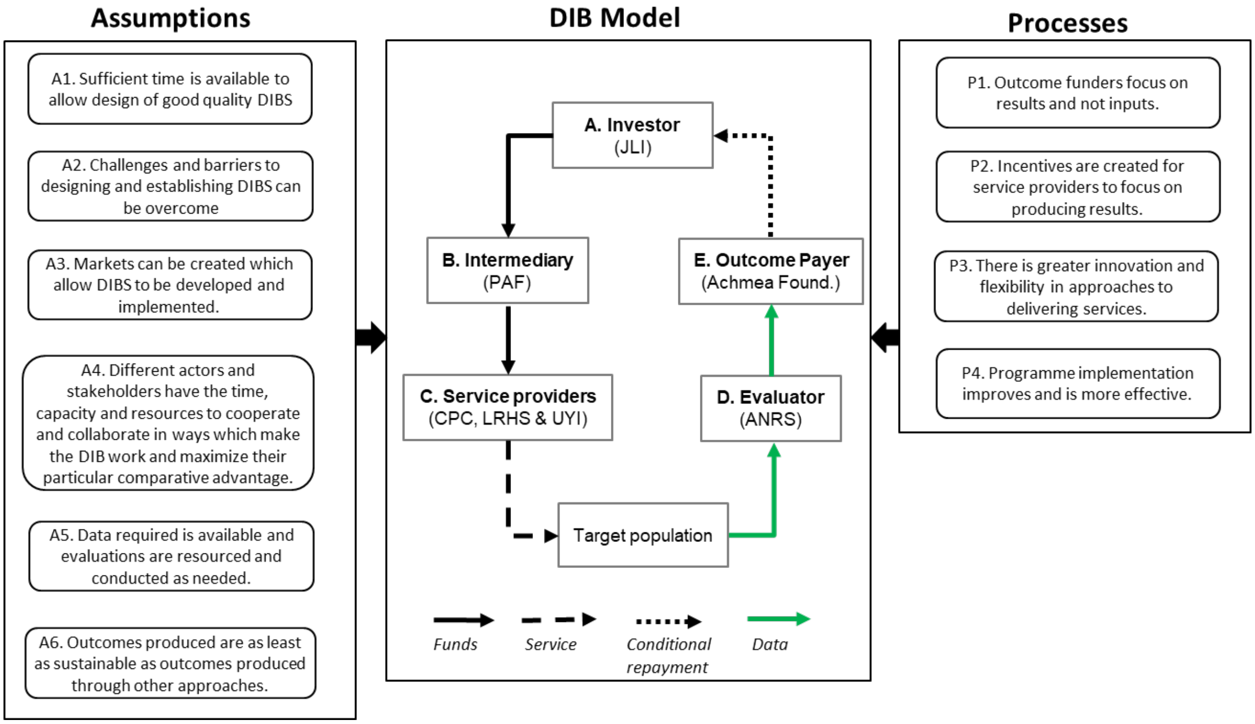

#### Appendix 3. Interviewed stakeholders

| Stakeholder | Role | Number of participants |
|---|---|---|
| Joep Lange Institute | Investor | 2 |
| Centre Pasteur Cameroon | Service provider | 2 |
| Laboratoire de Recherche sur les Hepatites virales et la Communication en Sante | Service provider | 5 |
| ANRS | Outcome evaluator | 1 |
| Legal department PharmAccess Foundation | Intermediary | 1 |
| PharmAccess Foundation | Intermediary | 3 |
| Medical advisor and country expert | External advisor | 1 |
| Achmea Foundation | Outcome payer | 3 |
| Global Loan Agency Services (GLAS) | Intermediary | 1 |
| Health economist expert | External advisor | 1 |
| Celle de PBF, Cameroonian government | Contextual actor | 1 |
| HEREG | External advisor | 1 |
| **Total** | **11 interviews** | **22 persons** |
